# Protocol for the Feasibility and Acceptability of Mentalization-based Treatment for Early Adolescents With Depression: A Short-Term Psychotherapy Approach for Patients and Families

**DOI:** 10.1101/2024.10.15.24315540

**Authors:** Javier Moran-Kneer, Javiera Duarte, Louise Alkabes-Esquenazi, Paula Solervicens, Francisca García, Marcelo Arancibia, Francisca Díaz, Nicole Avalos

## Abstract

**Background:** Approximately 13.4% of children and adolescents worldwide suffer from depression, with early onset often manifesting before the age of 14 and disproportionately affecting vulnerable groups. The prevalence of adolescent depression has increased since the COVID-19 pandemic, with significant long-term impacts on development, social functioning, and academic performance. Mentalization-based treatment (MBT), particularly the MBT-A model tailored for adolescents, has shown promise in addressing depression by increasing mentalizing capacity. This study explores the feasibility and acceptability of a short-term MBT-A intervention that incorporates both individual and family components to address depression in early adolescents.

**Methods:** This feasibility pilot study will recruit 15 adolescents aged 10--14 years who are diagnosed with mild to moderate depression from primary healthcare centers and psychological care clinics in Valparaíso, Chile. The participants will undergo a 12-session MBT-A intervention, which is structured into three phases: joint understanding of the problem, intensive promotion of mentalization, and evaluation of the process. Outcomes will be assessed through mixed methods, including quantitative measures (e.g., recruitment rate, adherence rate, effect size on depressive symptoms) and qualitative interviews with patients, families, and therapists. Discussion: Fostering mentalization in adolescents with depression may reduce depressive symptoms and enhance emotional regulation, benefiting both individuals and their families. This study aims to provide preliminary data to inform the planning of a larger randomized controlled trial (RCT). By incorporating family components, the intervention not only addresses individual symptoms but also promotes supportive family dynamics, which are crucial for sustaining therapeutic gains. This pilot study will identify practical aspects of implementing MBT-A and gather essential data on its feasibility and potential efficacy in a real-world setting.

## 1. Background and rationale

Approximately 13.4% of children and adolescents worldwide suffer from depression (1). Mental health issues often manifest before the age of 14 (2) and disproportionately affect vulnerable groups (3), with significant economic repercussions for public health that persist into adulthood (4). Adolescent depression, in particular, is a prevalent issue closely linked to self-harm, suicidal behaviors, and other risky behaviors (5). This condition frequently cooccurs with substance abuse, high-risk sexual behaviors, anxiety, and personality disorders (6, 7). Early onset of depression is tied to notable neurodevelopmental impairments during adolescence (8)). The prevalence of depression surged following the COVID-19 pandemic, with an estimated increase of 25% in depression and anxiety cases worldwide (9). International reports highlight four key areas impacted in children and adolescents: education, rights protection, poverty, and mental health (10, 11, 12).

Early adolescent depression, which occurs before the age of 15, is particularly significant because of its long-term impacts on development and functioning (13). Depression during these formative years is associated with impaired academic performance, which can have lasting effects on educational and occupational outcomes (14).

Early onset of depression can disrupt normal social and emotional development, leading to difficulties in forming and maintaining relationships, increasing the risk of social isolation (15) and increasing the risk of more severe and recurrent episodes in the future (16). Moreover, early adolescence serves as a critical period for brain development due to synaptic pruning— a crucial process for the maturation of regions involved in executive functions, emotional regulation, and social behavior (17). Disruptions in synaptic pruning can lead to maladaptive neural circuitry, which impacts emotional regulation and heightens the risk of depression (18). According to this evidence, addressing depression early in adolescence is crucial to mitigate these risks and support healthier developmental trajectories (19).

Importantly, these neural changes also impact adolescents’ capacity for mentalization—the ability to understand the mental states of oneself and others (20). Research has shown that the development and reorganization of brain areas during adolescence, particularly those involved in social cognition and executive functions, are crucial for the enhancement of mentalizing abilities (21,22). Recent studies have indicated that mental health disorders, including depression, are linked to dysfunctions in mentalizing capacities (23). Failures in mentalizing may play a crucial role in the development of depressive symptoms (24). During this time, adolescents must also develop new capabilities such as establishing relationships, gaining greater autonomy, and enhancing agency. These developments largely depend on the regulation of stress, mentalization, and reward systems. Consequently, poor mentalizing is linked to higher rates of depression during developmental transitions (25).

There is evidence suggesting that mentalization-based treatment (MBT), particularly when tailored for adolescents via the MBT-A model, is an effective intervention for addressing depression symptoms within this age group. A randomized controlled trial (26) revealed that short-term MBT-A therapy (12 sessions) reduced symptoms of depression and self-harm in adolescents more effectively than treatment as usual (TAU). Notably, 37% of patients in the MBT-A group no longer exhibited signs of clinical depression, whereas only 22% of patients in the TAU group did. The authors attributed this superiority of the MBT-A to improvements in adolescents’ mentalizing capacities and a reduction in their avoidant attachment behaviors. Similarly, a pilot randomized controlled trial (27) investigated an adaptation of the adult MBT introductory manual in a group format for adolescents. This study, which was conducted in a clinical setting, aimed to assess the feasibility and initial effects of a group-based Mentalization-Based Treatment for Adolescents (MBT-Ai). The findings indicated a reduction in self-harm behaviors among participants, supporting the potential of MBT-Ai as a promising intervention for this population.

In recent years, the focus of mentalization-based interventions has shifted from an individual approach to a family and community approach (28). These results also prove how intervention at the family level can promote mentalization and trust, even after the therapeutic intervention has been completed. This holistic approach not only fosters individual healing but also cultivates a supportive environment that perpetuates positive behaviors and emotional health across the community, thereby contributing to the sustainable well-being of all its members (29). Additionally, highlighting the role of attachment relationships in this model, it is important to note that the emotional and behavioral symptoms of children and adolescents appear to be positively related to parental well-being. Family relationships have been shown to be a protective factor in adolescent mental health (30, 31, 32). Conversely, evidence shows that poor family communication is associated with the development of psychopathology (33, 34). Adolescent depression is also associated with parental psychopathology and family dysfunction (35), which are factors that are reinforced by current sociohealth conditions (36). Parental involvement in psychotherapeutic interventions for adolescent depression enhances their effects, for example, through improved emotional coregulation, a key factor in treating these types of psychopathologies (37). Additionally, in the treatment of adolescents, parental emotional regulation skills and strategies should be addressed to reduce behaviors that could interfere with therapeutic processes (38).

In summary, we propose that mentalization-based psychotherapy, which includes both individual and family components, may serve as an effective intervention for addressing depression in early adolescents. This study presents the research protocol for a therapeutic model designed for this specific population. The theoretical foundation of this intervention is evolutionarily relevant, as it focuses primarily on strengthening psychologically vulnerable functions in the context of neurodevelopment. Moreover, it integrates the family as a central part of the treatment, an essential aspect considering its impact on the development and maintenance of psychopathology in adolescents and its protective value in this context. In this context, this therapy has a predominantly preventive nature, addressing depression at an early stage. This study provides both qualitative and quantitative data for the planning of a clinical trial to test the proposed hypotheses.

## 2. Methods/design

### 2.1. Aims

This feasibility pilot study aims to assess the feasibility and acceptability of short-term mentalization-based treatment for adolescents diagnosed with mild to moderate depression. The primary objectives include the estimation of a) the proportion of invited patients who agree to participate in the study (recruitment rate), b) the proportion of patients who complete the intervention relative to the initial number of participants (adherence rate), and c) the intervention’s effect size on depressive symptoms in adolescents. Additionally, on a qualitative level, the study aims to explore a) patient satisfaction with the intervention (acceptability) and b) the applicability of the treatment from the perspective of the therapists and supervisors involved in the intervention.

As secondary objectives, the study aims to determine the intervention’s effect size on a) depressive symptoms in other family members, b) anxious symptoms in all intervention participants, c) family cohesion, and d) overall adaptation of all family system members.

### 2.2. Design

The research corresponds to a pilot study aimed at evaluating the feasibility and acceptability of a brief family intervention informed by mentalization to address adolescents diagnosed with mild to moderate depression attending a secondary care center in the city of Valparaíso.

The mixed-methods study incorporates both qualitative and quantitative techniques for data collection and analysis to obtain and evaluate primary and secondary outcomes in the context of the intervention study. The design is based on the principles of the Consolidated Standards of Reporting Trials [CONSORT] - Extension to randomized pilot and feasibility trials (39).

According to the CONSORT, this type of design is recommended to evaluate the feasibility of interventions on a small and exploratory scale before scaling them up to randomized controlled trials (RCTs). This type of study allows for documenting changes to the original design, such as treatment type, eligibility criteria, and primary dependent variables, among others, providing reasons for these changes (40). Adicionalmente, el estudio sigue las guías de SPIRIT (41) (see Additional file 1)

### 2.3. Primary outcomes

The feasibility of the intervention will be assessed as follows:

a. Recruitment rate
b. Data attrition rate
c. Follow-up rate
d. Professionals’ reports who conducted the intervention through qualitative interviews.

The acceptability of the intervention will be evaluated as follows:

a. Adherence rate
b. Reports from adolescents and responsible adults through qualitative interviews and the Credibility/Expectancy Questionnaire (CEQ) (42).

### 2.4. Secondary outcomes

The following screening instruments will be applied to calculate the effect size of the intervention. The evaluation will be conducted on the responsible adult:

a. Depressive symptomatology was assessed via the PHQ-9 (43, 44).
b. Anxiety symptomatology according to the DASS-21 anxiety scale (45, 46)

The evaluation for adolescents will include the following:

a. Depressive symptomatology using the RCADS (47, 48, 49)
b. Externalizing and internalizing symptoms through the SDQ-SF (50).

Finally, at the family system level, family cohesion will be assessed via FACES III (51).

### 2.5. Additional assessments

At the psychotherapy process level, evaluations are carried out by external observers:

a. Therapeutic alliance through the VTAS-SF scale (52).
b. Adherence to the therapeutic model via the Mentalization-based Treatment Adherence and Competence Scale (MBT-ACS) (53).
c. Quality of patient mentalization based on the OMP-A (54)

Additionally, the progress of participants’ psychological well-being will be assessed through session-to-session evaluation via the CORE-OM (55) (for adults) and YP-CORE-OM (56) (for adolescents) scales.

The following table summarizes all the measures used in this study according to the different stages of the intervention:

**Table 1.** Overview of the assessments according to time points

|  | SCREENING | T0 | T1 | T2 | T3 |
| --- | --- | --- | --- | --- | --- |
| ENROLMENT |  |  |  |  |  |
| Eligibility screen | X |  |  |  |  |
| Informed consent | X |  |  |  |  |
| *Psychiatrist*<br>*assessment* | X |  |  |  |  |
| INTERVENTION |  |  |  |  |  |
| MBT intervention |  |  | X |  |  |
| ASSESMENTS |  |  |  |  |  |
| Adult | PHQ-9 | X |  | X | X |
|  | SDQ | X |  | X | X |
|  | FACES III | X |  | X | X |
|  | CEQ | X |  | X |  |
|  | DASS-21 | X |  | X | X |
|  | CORE-OM | X |  |  |  |
| Adolescent | RCADS-30 | X |  | X | X |
|  | YP-CORE-OM | X |  | X | X |
|  | CEQ |  |  | X |  |
| Observation<br>Instruments | OMP-A |  | X |  |  |
|  | MBT-ACS |  | X |  |  |
|  | VTAS-SF |  | X |  |  |

T0: Baseline, T1: Intervention, T2: Postintervention, T3: Follow-up, PHQ-9: Patient Health
Questionnaire, SDQ: Strengths and Difficulties Questionnaire, FACES III: Family

Adaptability and Cohesion Evaluation Scale, CEQ: Credibility/Expectancy Questionnaire DASS-21: Depression Anxiety Stress Scales, CORE-OM: Clinical Outcomes in Routine Evaluation – Outcome Measure, RCADS-30: Revised Child Anxiety and Depression Scale, YP-CORE-OM: Young Person-Clinical Outcomes in Routine Evaluation-Outcome Measure, OMP-A: Observing the Mentalization in Psychotherapy for Adolescents Scale, MBT-ACS: Mentalization-based Therapy Adherence and Competence Scale, VTAS-SF: Vanderbilt Therapeutic Alliance Scale.

**Figure 1:**
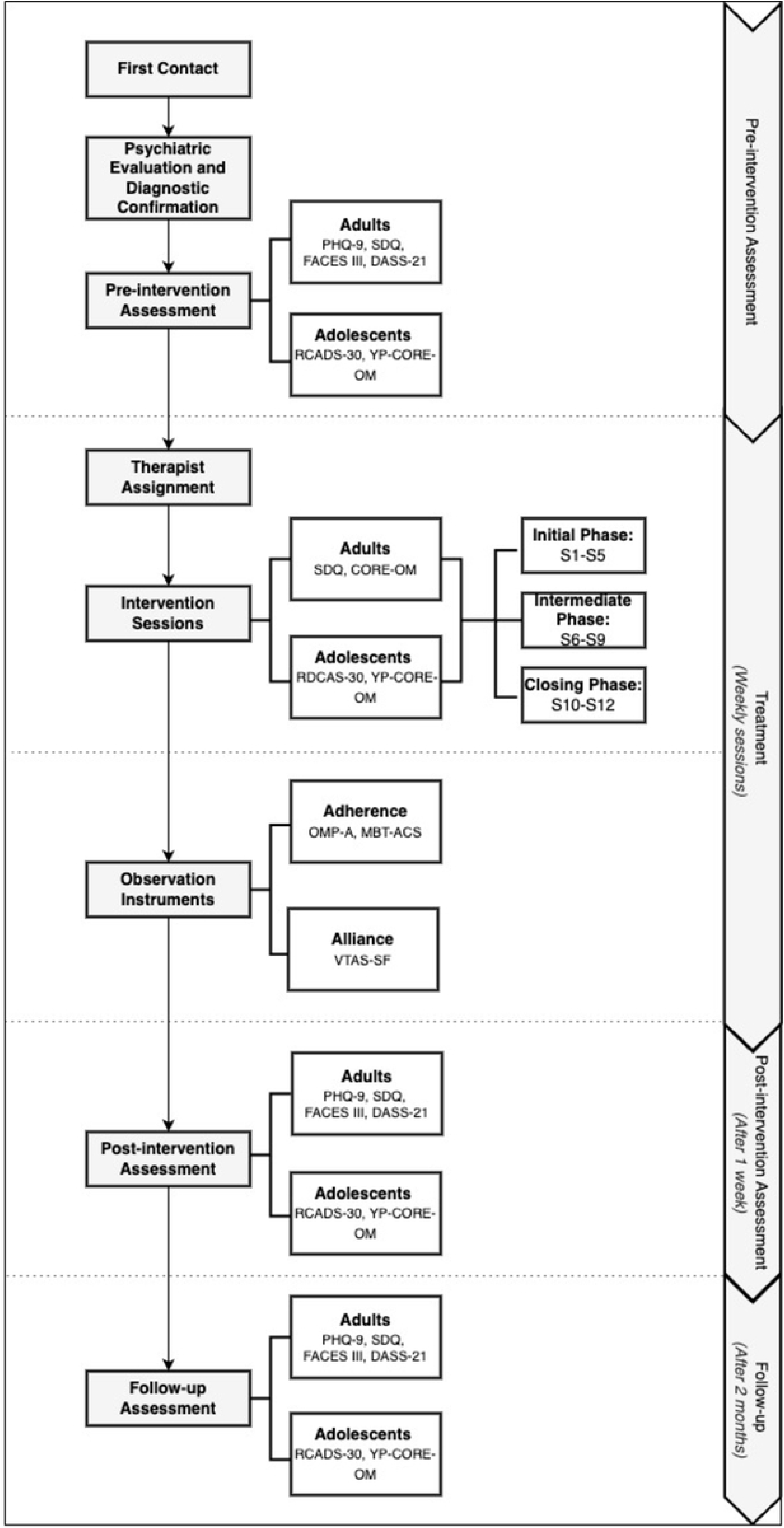

### 2.6. Participants

15 Adolescents aged between 10 and 14 years and diagnosed with mild to moderate major depressive disorder, according to the Diagnostic and Statistical Manual of Mental Disorders (DSM-5-TR) (57), will be recruited through direct referral from a primary healthcare center. Healthcare providers from this center will be responsible for conducting an initial screening of patients and referring them to the study until the total number of patients is reached. The mental health caregivers at the referral centers will consult with responsible adults of adolescents suspected of having depression regarding the possibility of being contacted by the research team to invite them to participate in the study. Informed consent will be obtained, and there will be no financial compensation for participation in the study. The psychological interventions will be provided in a teaching-assisted psychological care clinic.

Participant recruitment began on September 1, 2023, and will conclude on December 1, 2024. Follow-up of participants will continue until the end of 2024. The authors confirm that the ongoing trial for this intervention is registered at https://ichgcp.net/clinical-trials-registry/NCT06252090.

### 2.7. Eligibility criteria

Participants who agree to be contacted will be assessed by a psychiatrist, who will interview both the teenager and the responsible adult to confirm the diagnosis of mild to moderate depression. In addition to the diagnosis of depression, the inclusion criterion requires the presence of at least one legally responsible adult who consents to participate in the therapeutic process. The exclusion criteria included adolescents with diagnoses of autism spectrum disorders, psychosis, bipolar affective disorder, active suicidal ideation, or substance use disorder. Pregnancy is also an exclusion criterion. All participants must be users of the public healthcare system. The therapists include clinical psychologists and child and adolescent psychiatrists, who are part of the research team that contributed to the project’s formulation and will work in Child and Adolescent Psychiatric Services (CAPSI) specifically for this study. All the therapists had a minimum of 5 years of experience working with families and/or adolescents in the clinical setting.

The sample size calculation was initially based on an alpha value of .08, a 1-beta value of .05, and a large effect size for the depression outcome, anticipating significant clinical improvements due to the targeted nature of the MBT-A intervention. This assumption aligns with previous studies, such as Rossouw and Fonagy (26), where substantial effects were observed in similar populations. However, to ensure a robust design, we have also incorporated feasibility thresholds derived from the same study, setting recruitment at 60%, retention at 50%, and adherence at 70%. These thresholds provide a practical foundation for determining the sample size needed for a larger randomized controlled trial (RCT), ensuring that our estimates are grounded in both anticipated effect size and the practicalities of implementation in this specific population. With these parameters, a sample size of 15 families was estimated via G-Power 3.1.

### 2.8. Intervention

Short-term mentalization-based treatment for adolescents (MBT-A) with depression aims to increase the mentalization ability of adolescents to reduce depressive symptoms. This goal is achieved through both individual work and family intervention. This model retains the main characteristics of mentalization-based therapy, such as the therapist’s basic attitude toward actively promoting patients’ mentalization, the noncertainty of mental states, a focus on the here and now, and the use of affect as a mechanism for change. The treatment is structured approximately 12 weekly sessions organized into three phases. The model has a flexible structure, which is primarily based on achieving the objectives of each stage. The first phase’s main goal is the joint understanding of the problem, culminating in the creation of a written clinical formulation that is shared with the family between sessions 1 and 3. The second phase is characterized by intensive promotion of mentalization at both the individual and family levels, on the basis of the therapeutic focus established in stage 1. The third stage is oriented toward evaluating the process and establishing actions aimed at maintaining change. The treatment incorporates a psychoeducational component through a video animation introduced during the first phase. This component is designed to explain and discuss what depression is from a mentalization perspective, how it manifests in this stage, and the family’s role in treatment. Additionally, symptom tracking is incorporated throughout the process through routine outcome monitoring.

### 2.9. Treatment adherence

The interventions will be conducted by therapists trained in the MBT-A model (Mentalization-Based Therapy for Adolescents), who have completed 3 days of basic training provided by a certified instructor. All psychotherapy sessions will be video-recorded and evaluated by independent judges to determine adherence to the model via the MBT adherence and competence scale (53). Group supervision sessions will be held with the aim of reflecting on the development of clinical cases and promoting adherence to the model. Evaluations from independent judges will be integrated into weekly case discussions. Additionally, eight supervision sessions will be conducted with an expert supervisor in the MBT-A model throughout the study.

### 2.10. Data analysis

Descriptive analyses will be conducted to characterize the sociodemographic background of the sample, screening measures applied to participants, and primary quantitative feasibility outcomes (recruitment rate, data attrition rate, follow-up rate), as well as acceptability outcomes (adherence rate and quantitative evaluation of credibility/expectancy). For secondary variables related to participant symptomatology and family cohesion, multilevel analyses will be performed, considering the nesting of the therapists and the longitudinal nature of the data, to compare changes between the pre-assessment (T1), post-assessment (T2), and follow-up assessment (T3) periods. This analysis provides values for calculating effect sizes, a crucial input for determining the sample size needed for a larger-scale clinical trial. Finally, descriptive analyses will be conducted for additional assessments (therapeutic alliance, patient mentalization, treatment adherence, and progression of psychological well-being). All quantitative analyses will adhere to the per-protocol analysis principle (58) via Stata-18® software.

Qualitative data will be analyzed using thematic analysis (67) methodology. Interviews will be transcribed and coded to identify key themes, using NVivo software for data management. To ensure accuracy, findings will be triangulated across participants and validated through member checking The procedures associated with data management are described in the protocol approved by the Institutional Ethics Committee of the University of Valparaíso (CEC-UV 262-22).

## 3. Discussion

This protocol describes the process of a feasibility and acceptability study of short-term mentalization-based treatment for adolescents (MBT-A) diagnosed with mild to moderate depression. Adolescent depression requires not only symptomatic management but also consideration of neurodevelopmental changes during this stage. A mentalization-based approach strengthens structural functions (mentalization), which are crucial during adolescence for affect regulation. Promoting mentalization in individuals vulnerable to depression helps them cope with lifelong stressors, reducing the likelihood of relapse and even intergenerational transmission of depression (59).

There is also a need to incorporate the family as a key component, highlighting the impact of attachment relationships on the development of neurobiological structures involved in affect regulation and self-organization (60). Incorporating the family promotes the maintenance of changes and the prevention of future episodes (61).

Mentalization-based therapy, particularly its version for adolescents (MBT-A), has proven to be appropriate for treating depression in self-harming individuals (26). However, few studies on this therapeutic approach have focused specifically on adolescents with depression as the primary diagnosis, incorporating a family approach (62).

In accordance with the CONSORT statement (39), our main objective is to investigate the feasibility of an RCT in this group of patients, and as a secondary objective, to evaluate preliminary results of the intervention to assess possible treatment efficacy. The results from the interview analysis and the effect size calculation of the intervention for various outcomes are considered parameters and guidelines for a future RCT.

A pilot study is necessary before conducting an RCT in this group of patients, as depression has been reported as one of the diagnoses with the highest dropout rates in psychotherapeutic treatment, both before and during treatment, especially in the outpatient setting (63). There is also a need to maintain the adherence of participating family members.

The study design is adaptive, addressing specific concerns related to the depressive condition regarding the intervention or evaluations during treatment. Additionally, consulting adolescents’ oral evaluations at the end of treatment, along with quantitative data analysis, will further help develop and adapt the treatment in close collaboration with the adolescents.

Limitations of the study include the lack of a control group and potential recruitment biases due to high comorbidity with adolescent depression (64,65,66). Additionally, the intensity of weekly therapist supervision and differences in training levels are sources of bias in the results.

## Supplementary Information Acknowledgments

This work was possible thanks to the financial support of the National Fund for Health Research (FONIS), Chile. We would like to sincerely thank Dr. Efrain Bleiberg for his valuable suggestions and guidance throughout this project.

## Trial status

Currently, the clinical trial is in the recruitment phase. The intervention and follow-up of the participants are ongoing. The trial start date was May 5, 2023, and the intervention phase is expected to be completed in December 2024.

## Trial registration

Trial registration on ichgcp.net NCT06252090, February 6, 2024

## Harms

The procedures applied in this study are noninvasive, and no degree of discomfort or harm to the participants is expected. No ancillary or posttrial care or compensation in case of damage is planned.

## Data monitoring and auditing

There will not be a Data Monitoring Committee, as no harm is expected to result from study participation. Additionally, no auditing is conducted.

## Dissemination policy

The results of this research will be presented in publications, seminars and/or scientific and/or academic meetings.

## Abbreviations

CEQ: Credibility/Expectancy Questionnaire

CORE-OM: Clinical Outcomes in Routine Evaluations – Outcome Measures DASS-21: Depression Anxiety Stress Scale

DSM: Diagnostic and Statistical Manual for Mental Disorders FACES III: Family Adaptability and Cohesion Evaluation Scale III MBT: Mentalization-based treatment

MBT-A: Mentalization-based treatment for adolescents PHQ-9: Patient Health Questionnaire

RCADS-30: Revised Child Anxiety and Depression Scale-30 RCT: Randomized controlled trial

SDQ: Strengths and Difficulties Questionnaire

YP-CORE-OM: Young Person-Clinical Outcomes in Routine Evaluation-Outcome Measure VTAS-SF: Vanderbilt therapeutic alliance scale

## Authors’ contributions

JM and JD led the research project, performed project coordination, and provided overall supervision. JM, JD, JZ, FD and MA contributed to the conceptualization and design of the study. The content of the manuscript has been critically reviewed to ensure scientific integrity. JD, JZ, LE, MK, NA and FG contributed to the review of the literature and the objectives of the intervention. PS, SG, FD and NA contributed to the design and review of the manuscript writing. MA and FG conducted the discussion of the article. MA and JM contributed to the trial registration. Each author read and approved the final version of the manuscript, and all contributed significantly to the research and preparation of the article.

## Availability of data and materials

Data availability: The raw data supporting the conclusions of this article are accessible through a formal request process. Interested parties may submit requests for access to the data by contacting the corresponding author at. The procedures for reviewing and approving these requests will be outlined upon inquiry.

## Declarations

Ethics approval and consent to participate

This study was approved by the Scientific-Ethical Committee of the University of Valparaíso (CEC-UV), which guaranteed the informed consent of the participants and protected the confidentiality of the information collected. The study is registered with the number CEC-UV 262--22.

## Consent for publication

The participants had the right to know the results of this research. Informed consent will be obtained from at least one of the participants’ parents, as well as assent from the participant prior to inclusion.

## Contributor information

Javier Morán

Javiera Duarte, Johana Zapata

Louise Alkabes-Esquenazi

Silvia González, Paula Solervicens, Melanie Klagges

Francisca García, Marcelo Arancibia, Francisca Diaz

Nicole Avalos

## Recruitment status

Recruitment and intervention are ongoing

## Primary sponsor, principal investigator, and lead investigator

Javier Moran-Kneer is responsible for the design and conduct of the trial. He led the preparation of the protocol and revisions and the publication of the study results.

## Recruitment country

Chile

## Health conditions studied

Depression

## Intervention

Short-term mentalization-based treatment for adolescents (MBT-A)

## Key inclusion and exclusion criteria

Adolescents aged between 10 and 14 years and diagnosed with mild to moderate major depressive disorder according to the DSM-5-TR were included. Additionally, at least one legally responsible adult must consent to participate in the therapeutic process, and all participants must be users of the public healthcare system.

Excluded are adolescents with diagnoses of autism spectrum disorders, psychosis, bipolar affective disorder, active suicidal ideation, or substance use disorder. Pregnancy is also an exclusion criterion

## Study type

Feasibility and pilot study (single group)

## Date of first enrollment

01.09.2023

## Study status

The study is currently recruiting, intervening, and following up with patients.

## Primary outcomes

Feasibility of the MBT-A intervention (as indicated by recruitment rates, data attrition rates, follow-up rates, and qualitative interviews with professionals), acceptability of the intervention (as indicated by adherence rates, reports from adolescents and responsible adults through qualitative interviews and questionnaires, and necessary organizational resources (scientific personnel, recruitment networks, MBT-A training and supervision) to estimate the feasibility of an RCT.

## Secondary outcomes

Effect size of the intervention on depressive symptoms in adolescents, anxious symptoms in all intervention participants, family cohesion, and overall adaptation of all family system members.

## Protocol version

Version 1.0

## Data Availability

The raw data supporting the conclusions of this article are available upon request. Interested parties can submit data access requests by contacting the corresponding author at. The procedures for reviewing and approving these requests will include approval from the Institutional Ethics Committee of the University of Valparaíso.

